# Diagnostic accuracy of three mpox lateral flow assays for antigen detection in the Democratic Republic of Congo and the United Kingdom

**DOI:** 10.1101/2024.11.07.24316894

**Authors:** Elie Ishara-Nshombo, Anushri Somasundaran, Alessandra Romero-Ramirez, Konstantina Kontogianni, Daniel Mukadi-Bamuleka, Marithé Mukoka-Ntumba, Emile Muhindo-Milonde, Hugues Mirimo-Nguee, Jacob Parkes, Yusra Hussain, Susan Gould, Christopher T. Williams, Dominic Wooding, Juvenal Nkeramahame, Mikaela Watson, Hayley E Hardwick, Malcolm G Semple, J Kenneth Baillie, Jake Dunning, Thomas E Fletcher, Thomas Edwards, Devy M. Emperador, Hugo Kavunga-Membo, Ana Cubas-Atienzar

## Abstract

**Background:** The ongoing outbreaks of mpox, the disease caused by monkeypox virus (MPXV) highlight the urgent need for a rapid and low-cost MPXV diagnostic test to accurately detect and control mpox disease. In this study we assessed the diagnostic accuracy of three brands of rapid diagnostic tests for antigen detection (Ag–RDT) of MPXV.

**Methods:** We estimated the clinical diagnostic performance of FlowFlex Monkeypox Virus Antigen Rapid Test (ACON Biotech Co., Ltd., Hangzhou, China), Ecotest Monkeypox Antigen Rapid Test (Assure Tech. Co., Ltd, Hangzhou, China), and STANDARD Q Monkeypox Ag Test (SD Biosensor, Inc. Republic of Korea) using skin lesion swabs (SS) and upper–respiratory tract swabs (URS) from 68 participants in the Democratic Republic of the Congo (DRC) and from 16 in the United Kingdom (UK). The limit of detection (LOD) of the three Ag–RDT brands was evaluated using viral culture of the MPXV of the IIb lineage (EVAg, France).

**Findings:** Although the specificity of the three Ag–RDT was high (100%), sensitivity was estimated as 15·79% (95% CI, 5·52–37·57%) for Flowflex and Ecotest and as 10·53% (95% CI, 2·94–31·39%) for Standard–Q using SS in the DRC. The sensitivity was estimated as 0.00% (95% CI, 0·0–20·6%) among URS in the DRC. In the UK, the sensitivity of the three Ag-RDT was 0.00% among SS (95% CI, 0·0–12·7%) and among URS (95% CI, 0·0–21·5%). The LOD of all Ag–RDT was determined to be 1·0× 10^4^ pfu/ml (1·3×10^5^ copies/mL) using viral culture.

**Interpretation:** None of the three Ag–RDT reached the target clinical sensitivity and thus we do not recommend these as diagnostic or screening tool for suspected mpox cases. Marked improvement in sensitivity are needed for these Ag–RDT prior adoption. The recommended analytical LOD in the WHO target product profile (TPP) is at 10^6^ pfu/mL, being fulfilled by the three brands of Ag–RDTs suggesting that the analytical LOD stated in the TPP does not align with the desired clinical sensitivity. There remains an urgent need for accurate rapid Ag-RDTs for mpox diagnosis.

## Introduction

Mpox is a zoonotic disease caused by the monkeypox virus (MPXV) belonging to the *Orthopoxviridae* genus, which also includes the variola virus (smallpox). MPXV has two major clades: Clade I, subdivided in subclade Ia and Ib and Clade II, subdivided in subclade IIa and IIb.

Historically, mpox was an endemic disease of Central and West Africa. In May 2022, there was an upsurge of mpox cases with rapid expansion in non-endemic countries and it was declared the first mpox public health emergency of international concern (PHEIC) by the World Health Organization (WHO). Since then, over 100,000 cases of mpox with over 200 deaths have been described in more than 120 countries not previously considered endemic for mpox. The number of infections during the 20^th^ century has already been surpassed by cases occurring after the 2022 outbreak.^1^ On 14^th^ of August 2024, another mpox PHEIC was declared by WHO given the significant increase in mpox cases caused by the emergence of Clade Ib in the Democratic Republic of Congo (DRC) and neighbouring countries.^2^

In 2024, the Democratic Republic of the Congo (DRC), where mpox was first identified in 1970, reported the highest number of cases globally, with over 27,000 suspected infections and 800 deaths.^3^ In the United Kingdom (UK), most cases prior 2022 were associated with traveling from endemic countries. From August 2018 to September 2021, seven mpox cases were identified in the UK (four imported cases and three secondary). The discovery of the first mpox case of the 2022 global outbreak in the UK was on the 7^th^ May, a person who travelled from Nigeria, as of the 8^th^ of June, there were 336 laboratory-confirmed cases and 3,732 by end of 2022.

To confirm a clinical diagnosis, the WHO advises testing for MPXV as soon as possible in people who fit the suspected case definition. Laboratory–based real time PCR is the primary method used for MPXV detection. Laboratory–based PCR testing requires costly equipment, up front DNA extraction, and skilled personnel which may only be available in specialised laboratories haltering the rapid detection of cases during outbreaks. In contrast, rapid diagnostic tests (RDTs) are low cost, equipment– free, easy to use, suitable to use at the point–of–care (POC) and give results within 20 minutes.^4^ The utility of antigen RDTs for rapid detection of infected cases and patient isolation and management has been proven with many viral diseases, notably during the COVID–19 pandemic.^5^

The increasing global cases of mpox following the 2022 outbreak and the subsequent PHEIC 15 months later, brought to light the difficulties in meeting the increased demand of decentralised POC diagnostics for this highly infectious virus highlighting the urgent need for antigen RDTs (Ag–RDTs) for MPXV detection as a priority. This increased demand has resulted in dozens of these Ag–RDTs being available in the market. As of October 2024, there are 41 Ag–RDTs for MPXV at different stages of development of which 25 have achieved CE–IVD certification.^6^ Despite the increased number of Ag–RDTs for MPXV, clinical evaluation data is still lacking.^7^ To ensure reliable and accurate performance of Ag–RDTs, diagnostic evaluation studies across multiple, independent sites are required to generate the evidence based on their effectiveness, to guide implementation.

The aim of the study was to evaluate the diagnostic performance of three Ag–RDT brands for the detection of MPXV antigens: FlowFlex Monkeypox Virus Antigen Rapid Test (ACON Biotech Co., Ltd., Hangzhou, China), Ecotest Monkeypox Antigen Rapid Test (Assure Tech. Co., Ltd, Hangzhou, China), and STANDARD Q Monkeypox Ag Test (SD Biosensor, Inc. Republic of Korea) using skin lesion swabs (SS) and upper–respiratory tract swabs (URS) in two countries with different MPXV epidemiological characteristics, the DRC and the UK.

## Materials and methods

### Study settings and participants

In the DRC, individuals ≥ 2 years of age suspected to have mpox as per the WHO case definition^8^ were eligible to participate in the study. Eligible individuals were visited at home by a study health care worker to collect informed consent and collect the study samples. Participants were from the Maniema province and recruitment took place between January and December 2023. Paired SS and URS (upper respiratory swabs) were collected from all recruited participants in the study (n=68) and swabs were placed in 3mL of non-inactivating virus transport medium (VTM) (Jun Nuo, China). There were no follow–up visits and only skin lesion PCR results (considered the gold standard by the WHO) were provided to patients and used to make treatment decisions. Ethical approval was obtained by the DRC’s National Ethics and Health Committee (Comité National d’Ethique et de la Santé, CNES), reference 452/CNES/BN/PMMF/2023. All UTM samples were transported in cool boxes (2–8°C) to the National Institute for Biomedical Research (INRB) Biosafety Level 2 (BSL2) laboratories in Lubutu for processing and testing. All VTM tubes were processed within four hours for MPXV Ag–RDT testing and qPCR.

In the UK, retrospectively collected SS (n=30) and URS [n=23, (nasopharyngeal=1, oropharyngeal=22)] in universal transport media (UTM, RT-UTM Copan, Italy) from a cohort of 16 adult mpox patients enrolled at the Royal Liverpool University Hospital, Sheffield Teaching Hospital NHS Foundation Trust, and Royal Free London Hospital were used for this study. Patients were recruited during the last two outbreaks of mpox in the UK, 2018 and 2022. Patients were consented under the WHO ISARIC Clinical Characterisation Collaboration Protocol for severe emerging infections (ISRCTN66726260) ethical approval was obtained from the National Research Ethics Service and the Health Research Authority (IRAS ID:126600, REC 13/SC/0149). Mpox diagnosis was confirmed by sending samples to the UK Health Security Agency (UKHSA) for MPXV testing using qPCR. In addition to the samples from mpox positive patients, to fulfil with the minimum number of negative swab specimens for mpox diagnostic evaluations recommended by the FDA^9^, a set of 32 leftover nasopharyngeal samples in UTM (RT–UTM Copan) from prior COVID–19 studies^10^ were used as mpox negative controls. These were collected under the Facilitating Accelerated Clinical Validation of Novel diagnostics for COVID–19, and ethical approval was obtained from the National Research Ethics Service and the Health Research Authority (IRAS ID:28422, REC: 121 20/WA/0169). All samples were aliquots stored at –80°C and thawed for the first time for this study. Samples were processed and tested at the Biosafety Level 3 (BSL3) laboratories of the Liverpool School of Tropical Medicine (LSTM) as previously described.^11^

### MPXV Ag–RDT testing

In both sites, three Ag–RDTs were evaluated: FlowFlex Monkeypox Virus Antigen Rapid Test (FlowFlex hereinafter), Ecotest Monkeypox Antigen Rapid Test (Ecotest hereinafter), and the STANDARD Q Monkeypox Ag Test (Standard–Q, hereinafter). The three RDTs are based on immunochromatography and show the presence of MPXV A29L antigen using colloidal gold for visualization. Flowflex and Ecotest were CE in–vitro diagnostic (IVD) marked and commercially available while Standard–Q was for research use only (RUO) at the time of evaluation. All test brands can be used with SS; additionally, Flowflex can be used with serum, plasma and URS, Standard–Q in serum, plasma and whole blood and Ecotest in URS.

Test were performed in the laboratory in both countries (in INRB BSL2 laboratories in DRC and in LSTM BSL3 laboratories in the UK). Briefly, the specified amount of VTM/UTM confirmed by the manufacturers (200ul for Flowflex and Ecotest and 300ul for Standard–Q) was added into the extraction buffer and then, the number of drops of the extraction buffer specified in the instructions for use (IFU) was added into the sample well (four drops for Flowflex and Standard–Q and three drops for Ecotest). Test were read and interpreted visually after 15–30 minutes following the IFU. The results were read by two independent technicians and a third technician acted as a tiebreaker in case of discrepant results.

### Reference MPXV qPCR test

In both sites, DNA was extracted and a MPXV qPCR was performed using the same UTM/VTM tube used for the three Ag–RDT tests. In INRB, DNA was extracted from a 300ul aliquot of sample using the Natch 16S automated platform (Sansure Biotech, China) with the Nucleic Acid Extraction-Purification Kit (Sansure Biotech) according to the IFU. In LSTM, DNA was extracted from 200μL of UTM using the QiAamp96 Virus Qiacube HT kit (Qiagen, Germany) following the use IFU.

The same MPXV qPCR reference test was used in both sites, the Monkeypox virus Nucleic Acid Diagnostic Kit (Sansure Biotech). The qPCR was carried out using a MA-1620Q qPCR thermocycler (Sansure Biotech) at INRB and a QuantStudio 5 (ThermoFisher, USA) at LSTM. A qPCR result with a cycle threshold (Ct) ≤40 in the FAM channel (MPXV DNA) and Ct ≤40 in the Cy5 channel (internal control) was considered MPXV positive. This qPCR kit was used as the reference test as it has been successfully demonstrated to detect MPXV clades I, IIa and IIb^12^, it is CE IVD marked and has demonstrated higher diagnostic accuracy than other lab–based qPCR assays.^13^

### Analytical limit of detection (LOD) of the Ag–RDTs

A MPXV strain from the lineage II, B.1 (Slovenia_MPXV–1_2022) obtained from the European Virus Archive Global (EVAg, France) was cultured in Vero E6 cells (ECACC 85020206) in Dulbecco’s Modified Eagle Medium (Gibco, USA) plus 10% foetal bovine serum (Gibco) and 1% Penicillin/Streptomycin solution (Gibco) to generate the MPXV stock. A fresh aliquot was serially diluted ten–fold using UTM to produce concentrations from 1·0×10^4^ plaque forming units per mL (pfu/mL) to 1·0×10^0^ pfu/mL. The LOD was defined as the least concentrated dilution were all three replicates were positive. Quantification of the pfu/mL via plaque assay and the viral copy numbers per mL (copies/ml) of the serial dilutions and Ag–RDT testing to calculate the LOD were performed as previously described.^13^

### Statistical analysis

To assess the diagnostic accuracy of Ag–RDTs in patients with suspected mpox, point estimates of sensitivity and specificity were calculated for each Ag–RDT, based on results of the reference MPXV qPCR assay from the same VTM/UTM tube used for the Ag–RDT. The 95% confidence interval for each point estimate was derived based on Wilson’s score method. To compare performance of the Ag– RDTs at different Ct values, point estimates of sensitivity were stratified by Ct value of the reference test (all Ct ≤20, all Ct ≤25, all Ct ≤33, all Ct ≤40). Two-tailed Fisher’s exact and chi–square tests were used to determine non-random associations between categorical variables. Differences between the Ct values (expressed as mean± standard deviation (SD) in sample groups were assessed using the paired Student’s t–test. Statistical significance was set at α <0·05. Statistical analyses were performed using R scripts and GraphPad Prism 9·1·0 (GraphPad Software, Inc., CA).

## Results

### Clinical evaluation

In the DRC, a total of 68 participants were recruited for the study, of which 34/68 (50%) were male. The mean age of participants was 17 years (range 02–47 years). The median days from onset of symptoms was 4 (range 1–34). Fever (91%), skin lesions (100%), flu-like symptoms (75%), headaches (54%) and cough (50%) were the most prevalent symptoms (Table 1).

**Table 1.** Clinical characteristics of the recruited mpox patients from the DRC (n=68) and UK (n=16).

| Characteristics | DRC | UK |
| --- | --- | --- |
| Age [mean (min-max), N] | 17 (2–47), 68 | 35·1 (24–58), 16 |
| Gender [%M, (n/N)] | 50%, (34/68) | 100% (16/16) |
| Days from symptom onset [median (Q1-Q3); N] | 4 (3–7), 68 | 8 (4·25–12·75), 16 |
| Days 0–3 (n, %) | 32, 47% | 1, 6·25% |
| Days 4–7 (n, %) | 22, 32% | 6, 37·5% |
| Days 8+ (n, %) | 14, 21% | 9, 56·25% |
| **Symptoms** |  |  |
| Skin lesions | 100% (68/68) | 100% (16/16) |
| Fever | 91% (62/68) | 68·75% (11/16) |
| Flu like symptoms | 75% (51/68) | 25% (4/16) |
| Skin rashes | 0% (0/68) | 87·5% (14/16) |
| Headache | 54% (37/68) | 25% (4/16) |
| Cough | 50% (34/68) | 6·25% (1/16) |
| Sore throat | 37% (25/68) | 25% (4/16) |
| Nausea | 29% (20/68) | 0% (0/16) |
| Abdominal pain | 28% (19/68) | 0% (0/16) |
| Chest pain | 21% (14/68) | 0% (0/16) |
| Vomiting | 9% (6/68) | 0% (0/16) |
| Diarrhoea | 7% (5/68) | 6·25% (1/16) |
| Painful Urination | 6% (4/68) | 0% (0/16) |
| Eye discharge | 4% (3/68) | 0% (0/16) |
| Redness of eyes | 3% (2/68) | 0% (0/16) |

In the UK, 16/16 mpox patients (100%) were male with a mean age of 35·1 years (range 24–58 years). The median days from onset of symptoms was 8 (range 0–11). The most common symptoms were skin lesions (100%), skin rashes (87·5%) and fever (68·8%) (Table 1). Demographics and clinical characteristics of the negative cohort (COVID-19 patients) can be found in supplementary (S1).

In the DRC, 19/68 (28%) SS and 14/68 (21%) URS from individuals suspected of having mpox were positive when tested using the Sansure qPCR. Flowflex and Ecotest Ag–RDTs detected MPXV antigens in 3/19 MPXV positive SS, resulting in a clinical sensitivity of 15·79% (95% CI, 5·52– 37·57%), while Standard-Q detected MPXV antigen(s) in 2/19 resulting in a clinical sensitivity of 10·53% (95% CI, 2·94–31·39%). The Ag–RDT Flowflex was more sensitive when detecting MPXV antigen in SS with Ct ≤20 than those with Ct values ≥ 33 (P=0·008); however, this was not observed with the other Ag–RDT brands. None of the Ag–RDT brands detected MPXV antigen in URS, resulting in 0% (95% CI, 0–16·82%) sensitivity. The clinical specificity was 100% (95% CI, 92·73– 100%) for each of the Ag–RDTs performed in both sample types, SS and URS (Table 2).

**Table 2.** Clinical sensitivity and specificity of the MPXV Ag–RDTs Flowflex, Ecotest and Standard– Q using SS and URS from 68 mpox patients in the DRC compared to the Sansure qPCR.

| Sensitivity by Ct<br>(95% CI), N | SS |  |  | URS |  |  |
| --- | --- | --- | --- | --- | --- | --- |
|  | Ecotest | Flowflex | Standard-Q | Ecotest | Flowflex | Standard-Q |
| **Ct ≤20** | 0%<br>(0.0–79.4%), 1 | 100%<br>(20.7–100%), 1 | 0%<br>(0.0–79.4%), 1 | N/A | N/A | N/A |
| **Ct ≤25** | 14.29%<br>(2.6–51.3%), 7 | 28.57%<br>(8.2–64.1%), 7 | 14.29%<br>(2.6–51.3%), 7 | 0%<br>(0.0–79.4%), 1 | 0%<br>(0.0–79.4%), 1 | 0%<br>(0.0–79.4%), 1 |
| **Ct ≤33** | 27.27%<br>(9.8–56.7%), 11 | 27.27%<br>(9.8–56.7%), 11 | 18.18%<br>(5.1–47.7%), 11 | 0%<br>(0.0–39.0%), 6 | 0%<br>(0.0–39.0%), 6 | 0%<br>(0.0–39.0%), 6 |
| **Ct ≤40** | 15.79%<br>(5.5–37.6%), 19 | 15.79%<br>(5.5–37.6%), 19 | 10.53%<br>(2.9–31.4%), 19 | 0%<br>(0.0–21.5%), 14 | 0%<br>(0.0–21.5%), 14 | 0%<br>(0.0–21.5%), 14 |
| **Specificity %**<br>**(95% CI), N** | 100%<br>(92.7–100%), 49 | 100%<br>(92.7–100%), 49 | 100%<br>(92.7–100%), 49 | 100%<br>(90.8–100%), 54 | 100%<br>(90.8–100%), 54 | 100%<br>(90.8–100%), 54 |

In the UK, 16/23 URS (69·56%) and 27/30 SS (90%) from mpox patients were positive by Sansure qPCR. All 32 UTM samples analysed from the COVID–19 cohort tested negative for MPXV as expected. Zero positive results were obtained when testing either URS or SS regardless of the Ag– RDT brand used 0% (95% CI 0–20·59%). The difference in sensitivity in MPXV Ag–RDTs was significantly lower when using URS in comparison to SS (P value = 0·007). The specificity was 100% (95% CI 90·97–100%) for the three Ag–RDT brands on both sample types, SS and URS (Table 3).

**Table 3.**
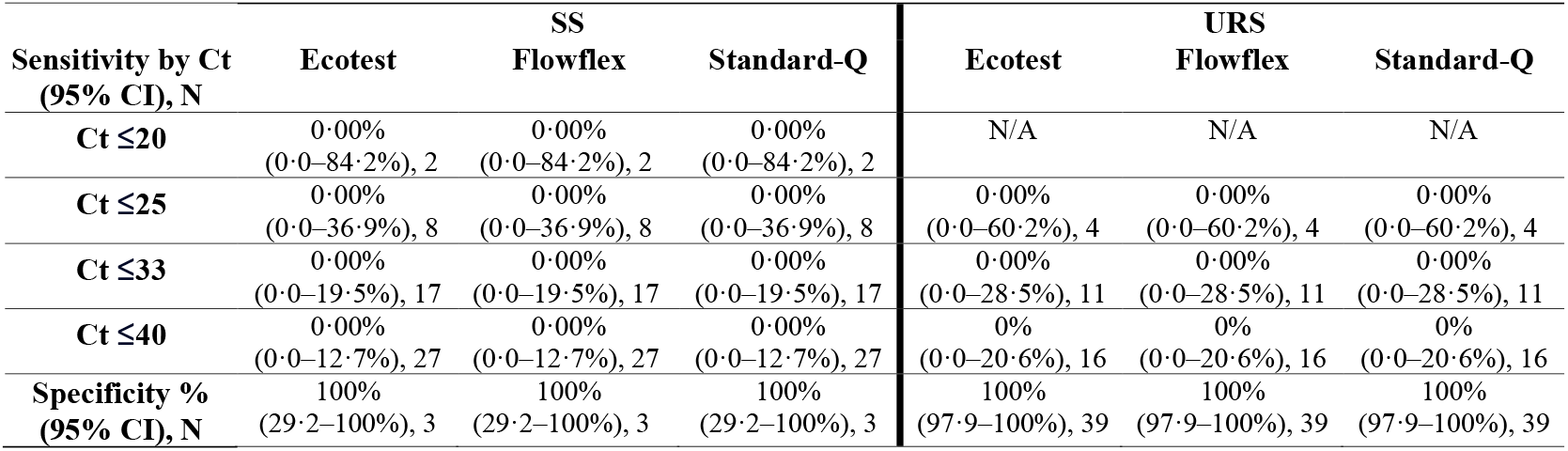
Clinical sensitivity and specificity of the MPXV Ag–RDTs Flowflex, Ecotest and Standard– Q using retrospectively collected samples: 30 SS and 23 URS from 16 mpox patients and 32 URS from 32 COVID–19 patients in the UK.

| Sensitivity by Ct<br>(95% CI), N | Ecotest | SS<br>Flowflex | Standard-Q | Ecotest | URS<br>Flowflex | Standard-Q |
| --- | --- | --- | --- | --- | --- | --- |
| **Ct ≤20** | 0.00%<br>(0.0–84.2%), 2 | 0.00%<br>(0.0–84.2%), 2 | 0.00%<br>(0.0–84.2%), 2 | N/A | N/A | N/A |
| **Ct ≤25** | 0.00%<br>(0.0–36.9%), 8 | 0.00%<br>(0.0–36.9%), 8 | 0.00%<br>(0.0–36.9%), 8 | 0.00%<br>(0.0–60.2%), 4 | 0.00%<br>(0.0–60.2%), 4 | 0.00%<br>(0.0–60.2%), 4 |
| **Ct ≤33** | 0.00%<br>(0.0–19.5%), 17 | 0.00%<br>(0.0–19.5%), 17 | 0.00%<br>(0.0–19.5%), 17 | 0.00%<br>(0.0–28.5%), 11 | 0.00%<br>(0.0–28.5%), 11 | 0.00%<br>(0.0–28.5%), 11 |
| **Ct ≤40** | 0.00%<br>(0.0–12.7%), 27 | 0.00%<br>(0.0–12.7%), 27 | 0.00%<br>(0.0–12.7%), 27 | 0%<br>(0.0–20.6%), 16 | 0%<br>(0.0–20.6%), 16 | 0%<br>(0.0–20.6%), 16 |
| **Specificity %**<br>**(95% CI), N** | 100%<br>(29.2–100%), 3 | 100%<br>(29.2–100%), 3 | 100%<br>(29.2–100%), 3 | 100%<br>(97.9–100%), 39 | 100%<br>(97.9–100%), 39 | 100%<br>(97.9–100%), 39 |

The comparison of the Ct values of paired URS and SS was assessed. There was a significant difference in Ct values between URS and SS sample groups (p-value=0·042) from DRC but not from the UK (Figure 1). The mean Ct values of the URS samples compared to the SS samples in the DRC were 30·7 (± 4·79) and 26·63 (±6·87), while in the UK were 27·2 (± 2·34) and 28·83 (±6·88), respectively.

**Figure 1.**
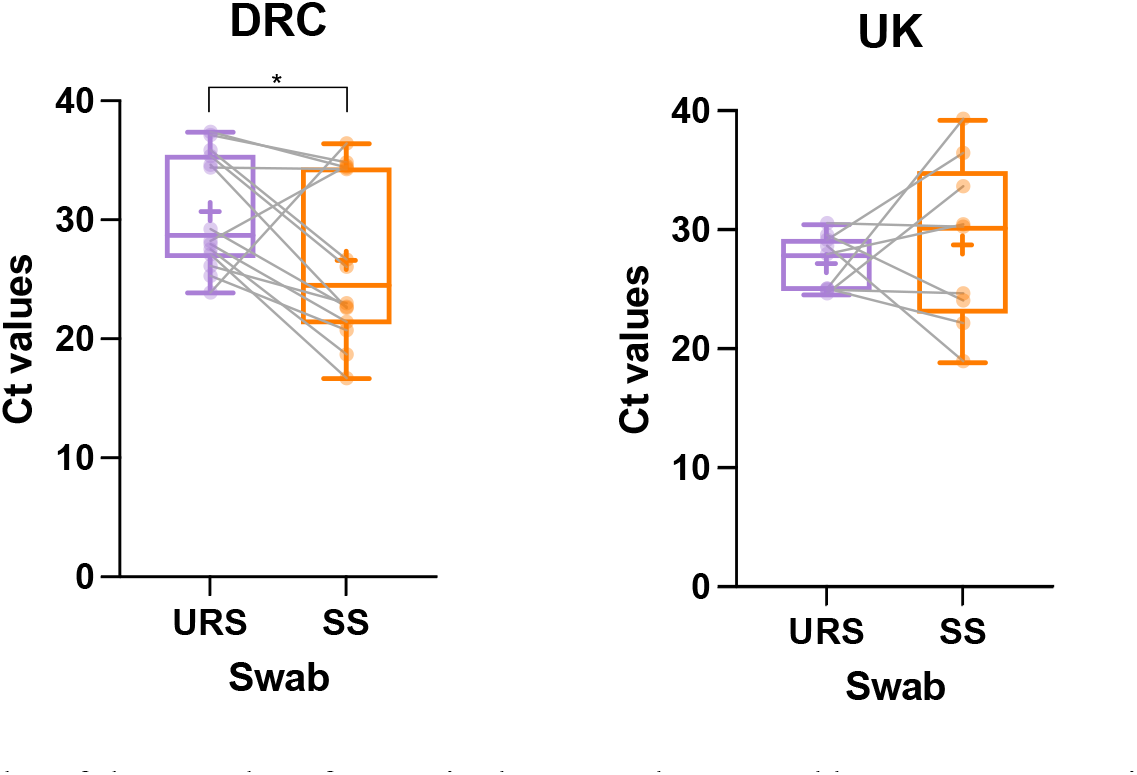
Boxplot of the Ct values from paired URS and SS tested by Sansure qPCR in DRC (n=14) and UK (n=9). The whiskers show the maximum and minimum values, and the vertical line represents the median. There was a significant difference (p–value <0·05) with higher Ct values in the URS group compared to the SS group in the DRC cohort.

No significant difference in sensitivity was found between the three Ag–RDT brands and between countries. The sensitivity of the Ag–RDTs was evaluated by onset of symptoms (Figure 2) with no significant differences observed on the Ag–RDT results and symptoms onset groups.

**Figure 2.**
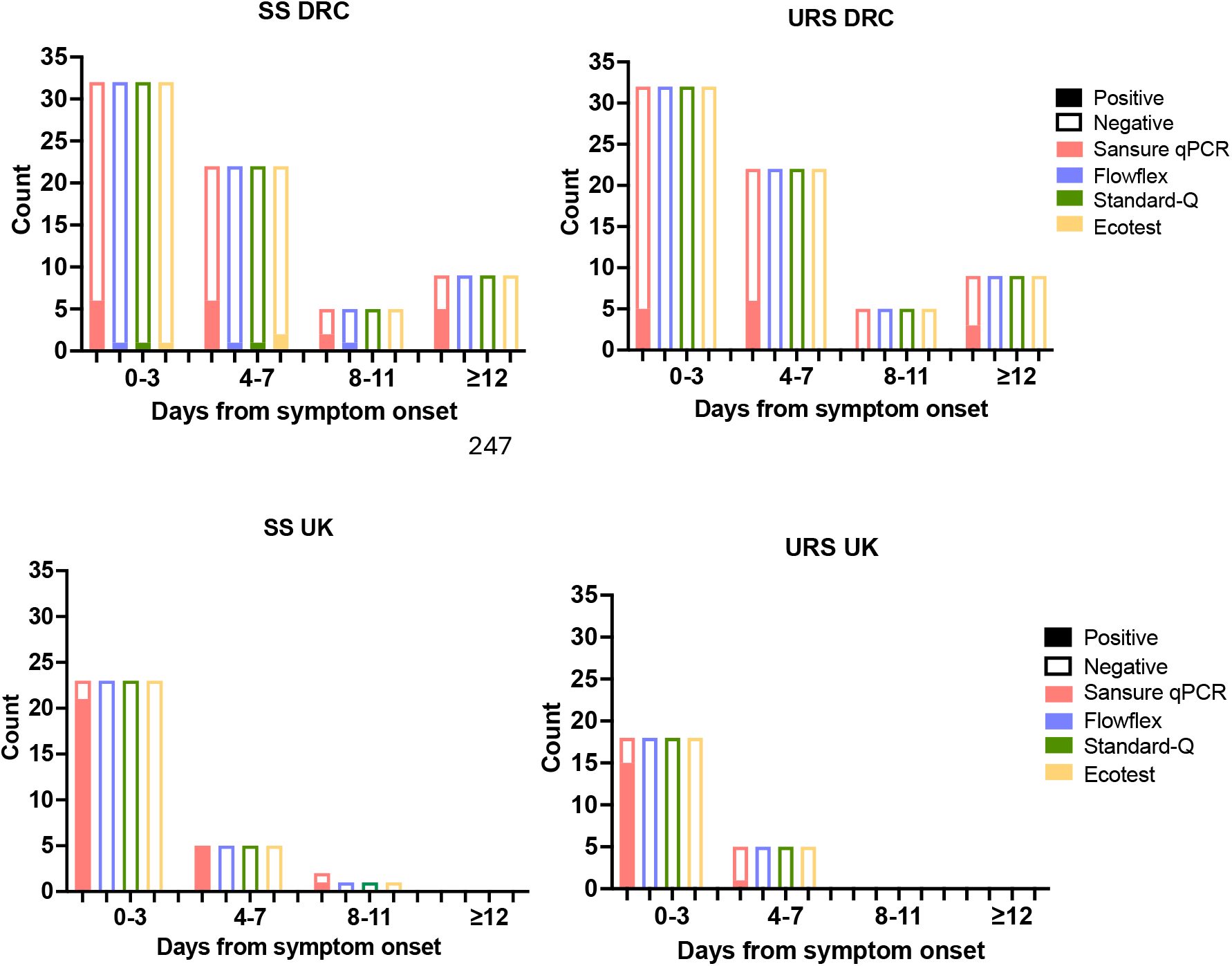
Number of positives and negatives results by test in SS and URS samples by days from symptom onset in the DRC and UK.

### Analytical Evaluation

The limit of detection of all the Ag–RDT brands using the MPXV viral stock was determined to be 1·0× 10^4^ pfu/ml (1·3×10^5^ copies/mL).

## Discussion

Following the recent PHEIC, WHO has issued is an urgent call to accelerate availability of POC diagnostics for mpox.^14^ The lack of validation data for MPXV Ag–RDTs represents a big gap in the diagnostics landscape that has slowed down rapid, effective responses to new outbreaks and ongoing endemic transmission.^15^ The primary aim of this study was to evaluate the diagnostic accuracy of three Ag–RDT brands Flowflex, Ecotest and Standard–Q using clinical samples from two different countries, DRC and UK.

WHO’s target product profile (TPP) for MPXV Ag–RDTs recommends minimal clinical sensitivity and specificity of 80% and 97% respectively.^16^ Specificity was fulfilled by the three Ag–RDT evaluated and both countries, but the sensitivity was extremely low, from 0% to 15·79%, for the three Ag–RDT brands in both countries, making these tests unsuitable for diagnostic use. Evaluation data on Ag–RDT for MPXV is very limited. A recent study using a Orthopoxvirus Ag–RDT prototype failed to detect MPXV antigens among 80 MPXV qPCR positive clinical samples in the UK (0% sensitive). The study suggested that the failure of detecting MPXV antigen in swab samples could be due to inhibition by the inactivating components of viral transport media (VTM), which can cause protein denaturation. In this study, we used non–inactivating swab transport mediums in both sites, and sensitivity was not improved. Nevertheless, the use of different types of swab transport mediums should be further investigated to optimise Ag–RDT performance including the use of dry swab to minimise the dilution effect. The dilution effect on Ag–RDT results for other viral diseases such as SARS–CoV–2 has been documented producing either lower sensitivity^17^ due to the additional dilution factor or false positives results due to non–specific electrostatic interactions between the antibodies in the assay.^18^

A study carried out prior the first global outbreak reported detection of MPXV antigens using the Ag– RDT Tetracore Orthopox BioThreat^®^ (Tetracore Inc. USA) in five out of the six tested MPXV positive samples with low Ct values (Ct=15–18 for Ag–RDT positives and Ct=22 Ag–RDT negative).^19^ However, this assay required sonication for swab material and dry ice/ethanol bath freezing followed by pestle grinding making it unsuitable for POC use.

Investigations of the analytical sensitivity of these Ag–RDT gave an LOD of 1·0 × 10^4^ pfu/mL, being more sensitive than previous analytical evaluations of Ag–RDTs for MPXV. The Orthopoxvirus Ag– RDT prototype had an LOD of 3·0 ×10^5^ pfu/ml^20^ and the commercially available Tetracore Orthopox BioThreat^®^ had an LOD of 1·5 × 10^6^ pfu/ml following sonication.^21^ The dilutions of the LOD experiment were made using the same UTM media used in the clinical samples tested in the UK and according with the results of the LOD (1·0 × 10^4^ pfu/mL/1·3×10^5^ copies/mL by qPCR), the Ag–RDTs should be sensitive enough to detect the clinical samples with Ct ≤ 28, which would yield a sensitivity of 57·8% and 35·7% in the DRC and 51·8% and 50% in the UK for SS and URS respectively. ^20^ These hypothetical clinical sensitivities would not fulfil current WHO TPP. The recommended analytical LOD in the WHO TPP is at 10^6^ pfu/mL, being fulfilled by the three brands of Ag–RDTs evaluated here and previously published, suggesting that the analytical LOD stated in the TPP does not align with the desired clinical sensitivity.

The use of LOD using viral isolates is often used as a proxy prior to having the test evaluated using clinical specimens;^17^ however in the present study and elsewhere^20^ the correlation between analytical and clinical sensitivity for MPXV has shown to be very poor, yielding lower sensitivity than expected among clinical samples. The reason for this great variability in antigen detection sensitivity between mpox clinical samples and mpox viral isolates is still uncertain. The quantity and type of accessible antigen in clinical samples needs to be further investigated. Additionally, the targeted antigens of the Ag–RDT evaluated in this study were A27L. Target antigens for other Ag–RDT brands include A27L, A35R, A5L, B6R, E8L, H3 and M1R.^6^ The antigen A27L has previously been suggested to be a good candidate as it is conserved and abundant within the virion; however the Ag–RDTS targeting this antigen in this study and reported elsewhere ^20^ did not yield acceptable sensitivity. This highlights the need for further evaluations using clinical samples with Ag–RDT targeting different antigen types.

The three Ag–RDT brands are designed to be used in other sample types besides SS, however none of the Ag–RDT detected MPXV antigens in URS, suggesting that the use of this sample type for antigen detection is not appropriate. One of the reasons for the lack of sensitivity of the Ag–RDTs in this sample type could be the lower viral loads compared to SS which was significant in the DRC cohort. Diagnostic evaluation studies using nucleic acid amplification found lower positivity rates in URS than SS^22,23^ which may be attributed to lower viral titres levels^24^ and/or earlier clearance in this sample type.^25^

In conclusion, the results of this study raise considerable doubts on the suitability of Ag–RDT for mpox surveillance and diagnosis, due to their poor clinical sensitivity. Serious validation should be performed regarding the reliability and accuracy of these devices before they are recommended for use in public health initiatives or surveillance studies. A second expression of interest has been launched by FIND following the recent PHEIC.^26^ Recommendations for next round of mpox Ag–RDT evaluations will include brands that detect different MPXV antigens, and evaluation of different swabbing techniques including the use of dry swabs to minimise dilution effect. The authors also recommend a revision of the analytical LOD for mpox Ag–RDTs as recommended in the WHO TPP as this does not correlate with the sensitivity on clinical specimens.

## Data Availability

All data produced in the present study are available upon reasonable request to the authors

## Author contributions

The study was planned and designed by AICA, JD, DME, HKM, EIN, DME and DMB. The Laboratory work was performed by AS, JP, YH, DW, NK, CTW, ARR and EMM under the supervision of AICA, TE, EIN and HKM. Data analysis and interpretation were conducted by AS, ARR, AICA, MW and EIN. AICA and EIN drafted the initial manuscript. The funding was obtained by AICA, HKM, RB, MGS, JKB, JD. Study oversight was provided by AICA and HKM. All authors reviewed and approved the final manuscript.

## Acknowledgements

We would like to thank the FIND team dedicated to managing and supervising this study: Emmanuel Agogo, Audrey Albertini, Daniel G. Bausch, Berra Erkosar, Susan Logoose, Michael Otieno and Aurelia Vessiere.

In DRC, we would like to thank all the participants who volunteered for this study. We would like to thank all the staff at Lubutu General Referral Hospital, especially Dr Jean Meniko, Dr Fiston Kaleba, Berthon Okoma, Naomie Koho, Emmanuel Matengano and Justin Kapela for their contribution to the success of this study. We would also like to thank Patrick Karungu for his work in entering the source data into the electronic database.

In the UK, we thank the ISARIC4C investigators [https://isaric4c.net/about/authors/], all the CRN, research nurses and clinical research fellows for supporting us with the sample collection and recruitment. We thank the CONDOR steering group: A. Joy Allen, Julian Braybrook, Richard Body, Peter Buckle, Eloise Cook, Paul Dark, Kerrie Davis, Gail Hayward, Adam Gordon, Anna Halstead, Charlotte Harden, Colette Inkson, Naoko Jones, William Jones, Dan Lasserson, Joseph Lee, Clare Lendrem, Andrew Lewington, Mary Logan, Massimo Micocci, Brian Nicholson, Rafael Perera-Salazar, Graham Prestwich, D. Ashley Price, Charles Reynard, Beverley Riley, John Simpson, Valerie Tate, Philip Turner, MarkWilcox, Melody Zhifang. We also thank the ISARIC CCP investigators: Dr Mike Beadsworth, Dr Ingeborg Welters, Dr Lance Turtle, Dr Jane Minton, Karl Ward, Dr Elinor Moore, Dr Elaine Hardy, Dr Mark Nelson, Dr David Brealey, Dr Ashley Price, Dr Brian Angus, Dr Graham Cooke and Dr Oliver Koch.

